# Human-Guided Agentic AI for End-to-End EHR Research: Sequential Antibody-Drug Conjugate Pathways in Breast Cancer

**DOI:** 10.64898/2026.10.13.26365479

**Authors:** Arturo Loaiza-Bonilla, Christos Colovos, Daniel Kim, James M. Grichnik, Kritika Singh, Paolo Tarantino

## Abstract

**PURPOSE:** To demonstrate an end-to-end research workflow combining a large electronic health record (EHR) resource and a multi-agent artificial intelligence (AI) system governed by Human-Guided Orchestration (HGO), using sequential sacituzumab govitecan (SG) and trastuzumab deruxtecan (T-DXd) orders in breast cancer as the clinical use case.

**METHODS:** A June 2026 EHR release contained 4.95 million patients, including 156,051 with breast cancer codes. Muse Spark supported cohort construction, executable statistical analysis, data-quality checks, literature retrieval, and reporting executed through Symbiotic Agent Teaming (SAT) under investigator direction. The main clinical measure was the second-drug record span, from first to last order. Analyses were descriptive and unadjusted; orders did not confirm administration or progression.

**RESULTS:** The workflow identified 106 patients: 73 had T-DXd ordered first and 33 had SG ordered first. Median second-drug spans were 63 days (interquartile range [IQR], 29–116) for SG after T-DXd and 49 days (IQR, 21–126) for T-DXd after SG; the median difference was −14 days (95% bootstrap CI, −41 to 21; distributional *P* = 0.42). Reported outputs additionally included cohort definitions, aggregate analyses, evidence context, figures, and a preliminary draft manuscript.

**CONCLUSION:** This study provides a concrete model for integrating large EHR resources, agentic AI, and human expertise across the research cycle. The ADC use case illustrates how computational execution and Expert-in-the-Loop (EITL) adjudication can produce interpretable research outputs and refine their scientific meaning, providing a foundation for extending and evaluating this approach across clinical questions.

**Context Summary:** *Key Objective:* How can clinical expertise and agentic AI be integrated from research-question formulation through EHR analysis and scientific reporting, using sequential SG and T-DXd orders as a clinical use case? Knowledge Generated
Muse Spark supported cohort construction, executable analysis, data-quality assessment, evidence retrieval, and reporting; investigators directed the work and adjudicated revisions. The resulting 106-patient cohort described recorded ADC pathways and the review determined record-span timelines.

## Introduction

Trastuzumab deruxtecan (T-DXd) and sacituzumab govitecan (SG) improve outcomes in overlapping populations with advanced breast cancer.^1–3^ T-DXd targets human epidermal growth factor receptor 2 (HER2), whereas SG targets trophoblast cell-surface antigen 2 (TROP2). Their payloads are different molecules that both inhibit topoisomerase I. Acquired alterations in the payload target and antigen can contribute to resistance, providing a biological rationale for studying sequential use.^4, 5^ These mechanisms do not establish which sequence benefits an individual patient.

Retrospective sequencing studies have reported limited clinical benefit from a second ADC, with substantial variation in receptor status, prior treatment, endpoint definitions, and selection.^6–10^ Electronic health record (EHR) studies already contribute to this evidence base.^8, 9^ A distinct measurement question is whether structured medication orders, without validated clinical abstraction, can describe the recorded sequence and span of use. An order may be planned, revised, duplicated, or canceled; it does not establish administration or progression.

Large EHR resources create opportunities to study clinical questions across longitudinal records, but doing so requires coordination of cohort definitions, executable analyses, data-quality review, evidence synthesis, and reporting. Agentic AI can connect these tasks through tools and iterative feedback. Recent work has applied this approach to EHR-informed clinical trial design.^11^ A complementary application is the production of an observational oncology study in which the clinical question, computational execution, and interpretation remain linked throughout the research process.

We conducted a working demonstration of this end-to-end EHR research using a formalized multi-tiered framework: Human-Guided Orchestration (HGO) for defining the clinical question and study parameters, Symbiotic Agent Teaming (SAT) for iterative code generation and evidence retrieval with Muse Spark, and Expert-in-the-Loop (EITL) intervention for clinical adjudication of data-quality assessments. Using sequential SG and T-DXd orders as the clinical use case, our objectives were to describe recorded ADC pathways and to document how this structured framework combined cohort construction, statistical execution, data-quality assessment, literature contextualization, and scientific reporting. End-to-end denotes the research cycle from question formulation through reporting. The methodological contribution is the coordination of these tasks and the investigator decisions that shaped the resulting evidence. We distinguish workflow implementation from comparative performance evaluation and retain established EHR data-quality principles.^12, 13^

## Methods

### Design and data source

This retrospective descriptive study used the Protégé research database (June 4, 2026 release), a longitudinal database that links de-identified electronic health records (EHR) and clinical notes with open medical and pharmacy claims. Data were de-identified under the Health Insurance Portability and Accountability Act (HIPAA) Expert Determination method. Protégé draws on routine-care EHR data from a network of 39 medium-to-large US health systems, aggregated by Loopback Analytics, and links each patient’s records to PurpleLab open claims through privacy-preserving tokens.

The release contains 4,950,000 patients residing across all 50 US states and the District of Columbia. Patients were selected based on documentation requirements (at least five clinical notes) and claims linkage, with enrichment for low-prevalence conditions; consequently, the database is not a probability sample. Overall, 99.4% of patients have linked medical or pharmacy claims, and subsets are additionally linked to radiology data and dated records of death.

The dataset encompasses 8.6 billion clinical events and 775 million clinical notes, with a median record span—from first to last dated clinical event—of 11.4 years (interquartile range, 7.0–18.8). Dated events fall predominantly between 2008 and 2024. For this study, we used data from 2020 to 2024, consistent with the dates of regulatory approval by the Food and Drug Administration (FDA) for both ADCs. Structured EHR data include demographics; encounters across ambulatory, telehealth, emergency, inpatient, and observation settings; diagnoses (predominantly ICD-10-CM); procedures (CPT, HCPCS, ICD-10-PCS, and local codes); laboratory results; vital signs; medication orders and fills; and surgical cases. Clinical notes retain their original text. Claims data include medical claim lines and headers with diagnosis and procedure codes and payer type, alongside pharmacy claims detailing National Drug Codes, days supply, and dispensing status.

Contributing site identities, specific geographic locations, and clinical settings of care outside participating organizations were unavailable. To protect privacy, dates were shifted by the data provider on a per-patient basis. Analyses therefore utilized within-patient intervals rather than comparisons of calendar dates across patients. The June 4, 2026, release date represents a data-processing date, not a common clinical follow-up boundary.

### Cohort and medication orders

Eligibility required an International Classification of Diseases, Tenth Revision, Clinical Modification (ICD-10-CM) C50-family diagnosis code and orders assigned to both drugs. The original case-insensitive name rule required both trastuzumab and deruxtecan for T-DXd, and sacituzumab or govitecan for SG. All order statuses were retained. Order start date was used, with record-creation date as the fallback. The earlier first qualifying order defined the group: T-DXd→SG or SG→T-DXd. See Methods S1-S6 for additional details.

Drug names were verified, and no changes to drug assignment, cohort membership, or reported results were required. Sacituzumab tirumotecan was absent from the data available at the database cutoff, and no clinical-trial participant data were included in the primary analysis. Verification found no brand-name omissions or shifted-date exclusions affecting the analytic cohort. Patients with the same first-order dates for both medications were excluded because sequence could not be determined. A metastatic diagnosis was not required. Operational details appear in the Data Supplement (Table S2).

### Endpoints and interpretation

The main endpoint was the second-drug record span: last qualifying order date minus first qualifying order date, with zero days for a single dated order. No dosing interval was added and no time-to-event censoring model was applied. The descriptive contrast was the difference in median spans for SG→T-DXd minus T-DXd→SG. It combines the identity of the second drug, treatment position, patient selection, and unmeasured tumor characteristics.

Six secondary comparisons were retained: first-drug record span; summed record span, adding the two ADC spans and the later systemic-therapy block; covered pathway span, from the first ADC order to the last systemic-therapy order; any further systemic therapy; number of distinct later agents; and interval from the last second-drug order to the latest dated record. The last measure is recorded observation after the second drug, not survival follow-up. The interval from the last first-drug order to the first second-drug order was descriptive and not tested. Further therapy required another systemic agent after the last second-drug order and could include continuing endocrine or HER2-directed therapy. Agent counts do not equal lines of therapy.

All spans include gaps between orders and may be truncated by the end of available records. They cannot identify progression, treatment cessation, or continuous exposure. Requiring records for both drugs selects on subsequent prescribing and continued data capture where a planned order does not establish survival to administration. Patients without a later second-drug order, including those still benefiting from the first drug or with death or loss of capture before a second order, could not enter. Overall survival was not analyzed since mortality ascertainment, treatment-initiation dates, and follow-up were not established as validated inputs to this analysis. If these inputs are validated, survival from second-ADC initiation could describe prognosis among patients reaching that treatment; it would not estimate the effect of choosing an ADC sequence at the first treatment.^14^

### Characteristics and data quality

Characteristics included age, recorded sex or gender, race, ethnicity, distinct previously ordered systemic agents, and C78-family and C79.3-family codes on or before the first ADC order. C78 records identify secondary malignancies of respiratory or digestive organs and do not capture all visceral disease. Demographic categories followed PCORnet Common Data Model version 6.0 in the analysis data. HER2 and hormone receptor status, disease stage, performance status, regimen-level treatment line, were not validated. Although the source included clinical notes, reasons for discontinuation were not established as validated analytic variables. A toxicity diagnosis or supportive-care order does not demonstrate that toxicity caused treatment cessation as attribution requires linking a documented treatment decision to the relevant ADC episode.

Data-quality assessment addressed conformance, completeness, and plausibility.^12, 13^ A secondary order-interval analysis used records meeting an order-density criterion described in the supplement (Table S6). This criterion did not confirm administration. Published clinical studies provided context, not validation of individual orders or equivalence of endpoints. Quantitative toxicity and laboratory-derived results are not reported because their ascertainment and release conditions were not adequately established.

### Statistical analysis

Continuous measures were summarized with medians and interquartile ranges (IQRs). Two-sided Mann–Whitney U tests compared distributions. Differences in medians had percentile bootstrap 95% CIs from 5,000 resamples; the reported seed was 7. A Mann–Whitney *P* value does not test the identical parameter as a CI for a median difference. Proportions had Wilson CIs, and the difference in proportions had a Newcombe CI. A pooled two-proportion *z* test compared further therapy. Holm adjustment covered the six secondary comparisons. Estimates were unadjusted because clinically essential covariates were not validated. The study used the available cohort, and uncertainty is conveyed by CIs rather than retrospective power calculations. Nonsignificant tests do not establish equivalence.

### Human-Guided Orchestration and Agentic Research Workflow

Muse Spark (Meta Superintelligence Labs) operated as an agent in a Symbiotic Agent Teaming (SAT) environment, utilizing tools for code execution in the licensed analysis environment, literature and registry retrieval, and file editing. It generated and ran cohort, endpoint, and statistical code; assessed data quality; retrieved evidence; prepared figures; and drafted manuscript text. Investigators provided Human-Guided Orchestration (HGO) by defining the clinical question and directing the analysis, while acting as an Expert-in-the-Loop (EITL) to review code and evidence, adjudicate proposed revisions, and edit all outputs as needed. Table 3 maps these tasks to human responsibilities and study outputs. Named reviewers, review scope, model and software versions, run dates, and available execution records are specified for completion in Methods S7.

Advisory critiques from AI agents and coauthor comments informed iterative revision, and the authors adjudicated changes. The original filtered time-on-treatment analysis was retired after EITL review. The final analysis used unfiltered first-to-last order spans. The original analyst confirmed that the reported medians, IQRs, confidence intervals, and statistical tests corresponded to this amended definition. The amendment and corresponding code and output identifiers are documented in Methods S6 and Appendix S1. Subsequent editorial review addressed literature, reporting, and aggregate consistency without accessing or rerunning patient-level data. Workflow outputs and revision decisions were described qualitatively. Research time and resource use were not prospectively measured, and no matched human-only workflow was evaluated. Consequently, no quantitative efficiency advantage is claimed. AI accuracy and independent reproducibility were not benchmarked.

### Ethics and reporting

The analysis used licensed deidentified records without patient contact. Bluebonnet Ethical Review served as the institutional review board of record. Its determination was that this activity does not meet the regulatory definition of human subjects research under the U.S. Department of Health and Human Services regulations at 45 CFR 46.102. To protect privacy, counts of 1–10 were withheld. Additional totals or counts were withheld when they would allow a hidden count to be calculated by subtraction. Reporting followed the STROBE and RECORD statements (Table S7).^15, 16^

## Results

### Selected cohort

Of 156,051 patients with a breast cancer code, 106 met the original analytic rules: 73 (69%) had T-DXd ordered first and 33 (31%) had SG ordered first. Cohort ascertainment and selection criteria are presented in Table S1. The breast-cancer-coded population was a search population, not a population established as eligible for both ADCs. Median age was 59 years (IQR, 51–66) and 55 years (IQR, 47–64), respectively. Both groups had a median of four previously ordered distinct systemic agents (Table 1). C78-family codes were recorded in 38 T-DXd→SG and 16 SG→T-DXd patients.

**Table 1.** Patient characteristics by first recorded ADC.

| Characteristic | T-DXd→SG<br>(*n* = 73) | SG→T-DXd<br>(*n* = 33) |
| --- | --- | --- |
| Age, years, median (IQR) | 59 (51–66) | 55 (47–64) |
| Race, White, No. (%) | 44 (60) | 20 (61) |
| Race, Asian, Black or African American, or other or unknown, No. (%) | 29 (40) | 13 (39) |
| Previously ordered distinct systemic agents, median (IQR) | 4 (2–6) | 4 (2–5) |
| C78-family diagnosis code, No. (%) | 38 (52) | 16 (48) |
| C79.3-family diagnosis code, No. (%) | 12 (16) | Suppressed |

Counts from 1 through 10 are suppressed. No row totals are added where they could reveal a suppressed cell. Race categories are combined only for disclosure control; the grouping is not a biological comparison. Overall race and ethnicity counts appear in Results. Prior agent count is not treatment line. C78 and C79.3 indicators do not capture all visceral or CNS metastases. IQR, interquartile range; SG, sacituzumab govitecan; T-DXd, trastuzumab deruxtecan.

Across the cohort, 14 patients were recorded as Asian, 12 as Black or African American, 64 as White, and 16 as other or unknown race. Ninety were recorded as not Hispanic and the remaining 16 combined Hispanic and other ethnicity categories and cannot be interpreted as a Hispanic-only count. Counts for selected small strata are suppressed. The data do not establish receptor status among patients in those strata.

### Record spans and later orders

Median second-drug spans were 63 days (IQR, 29–116) for SG after T-DXd and 49 days (IQR, 21–126) for T-DXd after SG (Table 2; Figures 1 and 2). The median difference was −14 days (95% CI, −41 to 21), with *P* = 0.42 for the distributional comparison. Median first-drug spans were 141 days (IQR, 71–334) and 189 days (IQR, 115–419), respectively (difference, 48 days; 95% CI, −33 to 200; *P* = 0.09; Holm-adjusted *P* = 0.54). These separate group medians do not estimate a within-patient change in treatment benefit.

**Table 2.** Main order spans and later systemic therapy.

| Measure | T-DXd→SG<br>(*n* = 73) | SG→T-DXd<br>(*n* = 33) | Difference<br>(95% CI) | *P* value |
| --- | --- | --- | --- | --- |
| Second-drug record span, days,<br>median (IQR) | 63 (29–116) | 49 (21–126) | −14 (−41 to 21) | 0.42 |
| First-drug record span, days,<br>median (IQR) | 141 (71–334) | 189 (115–419) | 48 (−33 to 200) | 0.09 |
| Interval between the drugs, days,<br>median (IQR) | 64 (30–161) | 40 (14–126) | — | — |
| Further systemic therapy, No. (%) | 47 (64) | 17 (52) | −13 (−32 to 7) | 0.21 |

Differences are SG→T-DXd minus T-DXd→SG. Span differences are in days; the later-therapy difference is in percentage points. Bootstrap CIs for median differences use 5,000 resamples. *P* values for spans are Mann–Whitney rank comparisons, not tests specifically of median differences.

**Table 3.** Human and agent contributions across the research workflow.

| Research stage | Workflow Tier | Model contribution | Human responsibility | Output in this study |
| --- | --- | --- | --- | --- |
| Question and specification | HGO | Translated the analysis specification into executable work. | Defined the clinical question and directed the analysis. | ADC order study with explicit eligibility and endpoints. |
| Cohort construction | SAT | Generated and ran selection code in the licensed environment. | Reviewed the analytic logic; confirmed drug-name and exclusion checks. | 106 patients with orders for both ADCs from 156,051 breast-cancer-coded patients. |
| Endpoint and statistical analysis | EITL | Generated and ran endpoint and statistical code. | Reviewed interpretation and adjudicated endpoint revisions. | Record spans, aggregate estimates, uncertainty, and comparisons. |
| Data quality and revision | EITL | Ran date and placeholder checks; supported advisory critiques. | Decided revisions in response to critiques and coauthor comments. | Amendment from filtered duration to record spans; explicit measurement limits. |
| Evidence context | SAT | Retrieved bibliographic and trial records and compared reported values. | Reviewed source support and clinical applicability. | Clinical context separating order spans from validated clinical endpoints. |
| Reporting and review | HGO/EITL | Generated figures, drafted text, and supported comment responses. | Reconciled critiques, edited drafts artifacts (manuscripts, tables, and figures) and retained responsibility for scientific claims. | Manuscript, supplement, figures, and documented revisions. |

The table summarizes tasks reported in the study methods, not a validated execution trace or performance comparison. Human reviewer identities and review scope require completion in Methods S7. The model supported the original analysis. Subsequent editorial and aggregate review did not access source records; it revised presentation and rebuilt the current figures and aggregate audit. The exact original orchestration, tool configuration, versions, and run dates should be documented from execution records. Abbreviations: Human-Guided Orchestration (HGO), Symbiotic Agent Teaming (SAT), and Expert-in-the-Loop (EITL) framework

**Figure 1.**
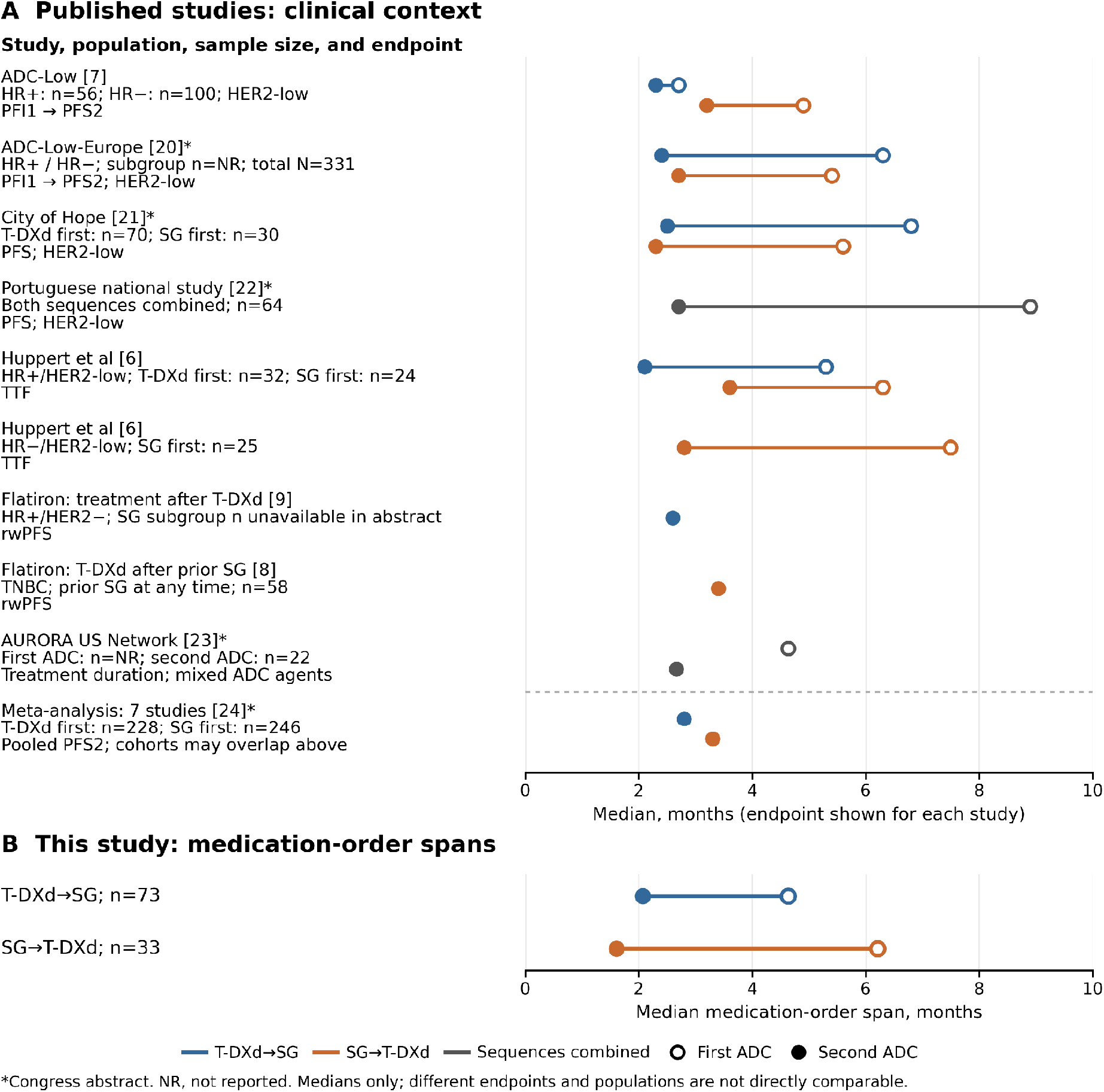
Published sequencing evidence and this EHR study. Open and filled circles show first- and second-ADC medians. Panel A presents published clinical endpoints; Panel B presents this study’s first-to-last order spans. Lines join estimates from the same subgroup, not individual patients. ADC-Low studies use first-ADC PFI and second-ADC PFS. AURORA points are unconnected because the first-ADC denominator is unspecified. Abstracts and pooled evidence are labeled; cohorts may overlap. Months (days/30.44) are display units, not harmonized endpoints. Confidence intervals are not displayed. Different populations and endpoints preclude effectiveness comparisons or validation of order spans. Sources, qualifications, and abbreviations are in Table S5.^6–9, 20–24^

**Figure 2.**
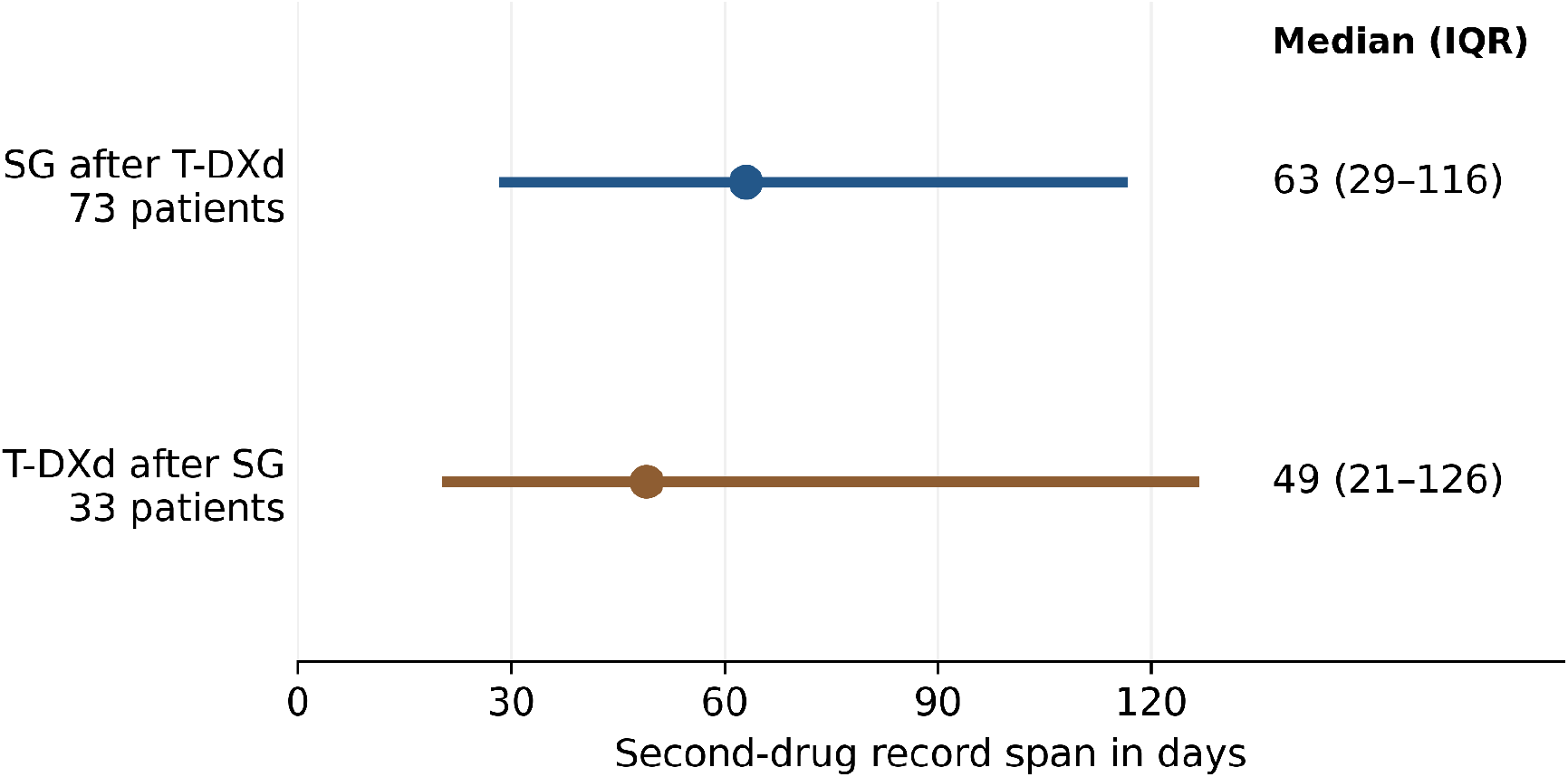
Second drug record span by first recorded ADC. Points show group medians and horizontal segments show interquartile ranges, not confidence intervals. The second drug is SG in the T-DXd-first group and T-DXd in the SG-first group. Each span runs from the first to last recorded order for that drug. These are not progression-free survival, treatment-discontinuation, or administration-confirmed duration estimates. Figure values reproduce the reported aggregates; the source cohort has not been rerun during this revision.

The median interdrug interval was 64 days (IQR, 30–161) for T-DXd→SG and 40 days (IQR, 14–126) for SG→T-DXd. Median summed spans were 287 and 393 days, and covered pathway spans were 426 and 514 days. None of the six secondary comparisons had a Holm-adjusted *P* value below 0.05. Full secondary estimates and their definitions appear in the supplement (Table S3).

Further systemic therapy was recorded in 47 of 73 patients (64%; 95% CI, 53%–74%) and 17 of 33 patients (52%; 95% CI, 35%–67%). The difference was −13 percentage points (95% CI, −32 to 7; *P* = 0.21; Holm-adjusted *P* = 0.84). Eribulin was recorded in 24 and 12 patients, respectively. Mean numbers of distinct later agents were 1.45 and 1.24. Median recorded observation after the second drug was 154 and 184 days. These intervals affect the opportunity to observe subsequent orders and do not establish vital status.

### Order interval assessment

Among order series meeting the secondary density criterion, median per-patient mean intervals were 21.7 days (IQR, 11.0–28.2) for T-DXd and 12.0 days (IQR, 7.0–17.0) for SG (Table S4). The distributions reflect different order patterns but do not validate administration. The T-DXd lower quartile was substantially shorter than its 21-day dosing schedule, consistent with possible duplicate or rescheduled orders. SG’s days 1 and 8 schedule creates alternating 7- and 14-day intervals but an average interval alone cannot establish adherence.^17, 18^

### Workflow outputs and scientific revisions

The reported workflow produced cohort and endpoint code, aggregate analyses, data-quality assessments, a clinical evidence comparison, figures, and manuscript text (Table 3). Expert-in-the-Loop (EITL) review changed the analysis and its interpretation: the AI-proposed filtered duration endpoint was replaced with descriptive first-to-last order spans, and clinical labels were excluded because their validity had not been established. This episode illustrates how expert adjudication can identify a mismatch between computable variables and clinical meaning and revise the resulting scientific claims. Subsequent editorial reconciliation clarified the descriptive interpretation and removed unsupported quantitative toxicity and laboratory results. Author verification of medication names and exclusions left cohort membership and numerical results unchanged. These observations document completed tasks and revision decisions, rather than comparative AI performance.

## Discussion

The principal contribution is a worked example of human-guided agentic AI spanning the EHR research cycle. Muse supported computational execution and reporting, while investigators supplied clinical direction and adjudicated scientific revisions. Applied within a 4.95-million-patient resource, the workflow identified 106 patients with orders for both ADCs, T-DXd was recorded first in approximately two thirds, and median second-drug spans were 63 days for SG and 49 days for T-DXd. The study contributes a description of recorded ADC pathways and an operational account of integrating clinical expertise with agentic execution across cohort construction, analysis, evidence synthesis, and reporting (via HGO, SAT, and EITL). Its methodological contribution is the explicit connection between computational outputs, expert scientific judgment, and revision of the resulting claims.

The methodological contribution lies in connecting research stages and making scientific decisions visible. The endpoint amendment illustrates why this matters: executable code can produce numerical results even when the intended clinical interpretation exceeds what the records establish. Review led to a measure aligned with the observable data and a corresponding revision of the claims. Alongside prior agentic work on EHR-informed trial design,^11^ this oncology use case illustrates the production and critical refinement of an observational study. Performance of transferable elements including the division of labor, explicit analytic definitions, and record of accepted revisions remain to be tested.

Clinical sequencing cohorts provide relevant context but measure different outcomes. ADC-Low reported short second-ADC PFS, and Huppert and colleagues reported receptor-specific time to treatment failure.^6, 7^ Flatiron-based studies used curated real-world progression outcomes.^8, 9^ Figure 1 presents this clinical context alongside additional published cohorts and pooled evidence, with the present order spans in a separate panel. Table S5 documents the populations, endpoints, estimates, and source qualifications. The comparison shows where this EHR study sits within the sequencing literature and identifies the clinical information needed to connect order-based research with outcome-based studies (Methods S5), A first-to-last order span omits the interval following the final recorded order and lacks a progression or discontinuation event. Numerical differences from published PFS or time to treatment failure therefore cannot be interpreted as differences in clinical benefit. Similar medians would not establish criterion validity.

The observed group difference cannot guide sequencing. Orders for both drugs did not establish that both were appropriate alternatives at the same disease state or treatment line. Joint eligibility at the first ADC was not established, and clinically essential covariates were not validated. No adjusted sequence-effect estimate is reported. Conditioning on both drugs selects for subsequent prescribing and continued data capture. The numerically shorter span with T-DXd second does not contradict studies reporting longer PFS with that order. However, the endpoint and population differ. Conversely, a nonsignificant comparison provides no evidence that the sequences are clinically equivalent. The unequal group sizes may reflect tumor phenotype, prior therapy, prescribing eligibility, or data capture. Equal median numbers of prior agents do not imply equal treatment lines. Shifted calendar dates also limit interpretation of prescribing-era differences.

Biological cross-resistance is plausible because the two payloads inhibit topoisomerase I, and acquired TOP1 and antigen alterations have been documented.^4, 5^ This cohort cannot test that mechanism. A shorter second-drug record span could reflect later disease, clinical deterioration, toxicity, incomplete records, shorter observation, or ordering practices. It should not be interpreted as proof of resistance or diminished efficacy.

Verification resolved the medication-name and exclusion concerns without changing the analytic cohort or results. Tirumotecan was absent from the source data, no clinical-trial participant data entered the primary analysis, and no brand-name omissions or shifted-date exclusions affected cohort membership. Remaining measurement limitations concern what an order represents: all-status records may include therapy never administered, repeated or planned orders can alter spans, and untied placeholder dates were not systematically screened. Agreement between aggregate order intervals and a known dosing schedule supports plausibility but does not validate administration or clinical endpoints.

The next opportunity is to evaluate whether this division of work can make medical research more scalable and iterative. Larger, clinically curated datasets could extend this work to validated receptor status, treatment episodes, progression, mortality, and reasons for discontinuation. Greater scale can improve precision, but curation, appropriate study design, and direct workflow evaluation are needed to establish clinical validity and research performance. This study did not measure research time, cost, error rates, independent reproducibility, or transfer across datasets. Future evaluations should compare complete workflows using matched questions, independently checked source records and analyses, and prospectively recorded resource use. Oncology curation programs such as Project GENIE provide relevant experience in source-record review and human quality assurance.^19^ Such evaluation would establish when agentic execution meaningfully extends investigators’ research capacity.

Clinical interpretation is chiefly limited by selection and measurement: patients were selected for orders for both ADCs, clinical outcomes were not validated, and receptor status, treatment line, and performance status were not established as reliable analytic variables. These features may exist in source records. The spans lack a validated discontinuation event and do not address administrative truncation through survival methods. Database size does not establish representativeness or compensate for selection and measurement error. Available sample size limits precision, and small-cell restrictions constrain subgroup reporting. Clinical toxicity cannot be inferred from unvalidated diagnosis codes, supportive-care orders, or laboratory record counts.

This study provides a worked example of end-to-end medical research combining a large EHR resource, the model, and human expertise. The workflow connected a clinical question to cohort construction, executable analysis, data-quality assessment, evidence synthesis, and scientific reporting. Applied to ADC sequencing, it characterized recorded treatment pathways and showed how EITL review shaped endpoint selection and interpretation. Its broader contribution is a concrete research model detailing how to implement Human-Guided Orchestration (HGO) across a multi-agent system. By isolating execution tasks to Symbiotic Agent Teaming (SAT) and clinical interpretation and adjudication to EITL checkpoints, this demonstration provides a scalable foundation for testing whether formalized human-agent architectures can support and accelerate accurate, reproducible, and efficient evidence generation across clinical questions and larger, clinically curated datasets. Prospective evaluation of analytical accuracy, reproducibility, research time, and expert effort would establish when human-guided agentic workflows can expand the scope and pace of clinically consequential research.

## Data and code availability

Patient-level records are licensed third-party data and are not redistributed with this manuscript. The accompanying files contain reported aggregate results, operational definitions, and code for a limited audit of publicly reported counts and figure generation. That code does not reproduce cohort selection, patient-level statistics, or clinical validation. No public repository DOI or independent reproduction is claimed.

## Declarations

### Ethics approval and consent to participate

This study was conducted in accordance with guidelines of the Declaration of Helsinki, Belmont report, and U.S. Common rule. In keeping with 45 CFR 46.104(d)(4), this study was performed utilizing retrospective, deidentified clinical data, and is considered IRB exempt with waivers of patient informed consent. Waiver of patient consent and exempt status was determined by Bluebonnet IRB. **Consent for publication:** Not applicable.

## Acknowledgments and AI disclosure

This study was supported through using an internal Meta scaffold around Muse Spark 1.3 and an internal version of the next generation of Muse models which assisted the authors in all the stages of the study supporting the original computational and writing workflow. Advisory agent critiques and coauthor review informed the scientific revisions. The authors adjudicated all recommendations. No AI system is an author. Authors remain responsible for the accuracy of all methods, results, references, and declarations.

