## Supplemental Files for "Human-Guided Agentic AI for End-to-End EHR Research: Sequential Antibody-Drug Conjugate Pathways in Breast Cancer"

### Data Supplement

This supplement documents the clinical use case and the formalized multi-tiered research framework—incorporating Human-Guided Orchestration (HGO), Symbiotic Agent Teaming (SAT), and Expert-in-the-Loop (EITL) oversight. It includes operational definitions, the architectural division of work, analysis amendments, and reproducibility information. Values are retained from the reported aggregate results unless explicitly described as an arithmetic check. Author verification of drug identity and exclusions required no change to the cohort or results. The editorial revision did not independently rerun patient-level analyses.

### Methods S1 Data source and cohort construction

The study used a deidentified longitudinal electronic health record (EHR) database containing 4,950,000 patients in its June 4, 2026 release. Available domains included diagnoses, medication orders, procedures, demographics, clinical notes and other dated records. The identities, geographic distribution and clinical settings of contributing organizations were not available to the investigators, and care delivered outside those organizations was not captured. The release date is a database version identifier, not the end of a patient's clinical follow-up. The provider shifted clinical dates within patients. Preservation of the intervals used in this analysis depends on consistent date transformations across the relevant event tables.

Eligibility required at least one ICD-10-CM diagnosis in the C50 family, at any recorded date, and medication orders matching both study-drug definitions. Metastatic disease, a particular receptor subtype and a minimum prior observation period were not required. The search population therefore included patients who may not have been eligible for either or both drugs as it is not an appropriate denominator for estimating uptake of sequential ADC treatment.

Orders were identified from case-insensitive source medication names. T-DXd required both 'trastuzumab' and 'deruxtecan'. SG required 'sacituzumab' or 'govitecan'. Drug-name verification found no incorrect assignments and required no change to cohort membership or reported results. Sacituzumab tirumotecan was absent from the data available at the database cutoff. No clinical-trial participant data were included in the primary analysis. No brand-name omissions affecting cohort membership were found. This source-name verification does not establish drug administration. No order-status filter was applied. Each order was dated using its start date or, if that was missing, its record-creation date as these fields can represent planned rather than administered treatment.

The drug with the earlier first matching order defined the sequence label. Patients with identical first-order dates were excluded because ordering could not be resolved and the source analysis characterized those tied dates as placeholders. Other placeholder dates were not systematically screened. A valid-calendar-date check excluded no patient. The initial draft raised concern about an exclusion based on shifted dates relative to the release date. Subsequent author verification found no shifted-date exclusions affecting the analytic cohort and no change to the reported results. Fewer than 11 patients were excluded overall, and exact small exclusion counts are withheld. The final reported cohort contained 106 patients: 73 with T-DXd ordered first and 33 with SG ordered first. The reported aggregates remain unchanged after author verification. The editorial revision did not independently rerun patient-level cohort selection.

Medication orders establish recorded prescribing activity, not dispensing, administration, adherence, continuous exposure or treatment indication. Sequence labels refer to first-observed ordering and do not establish consecutive lines of therapy, absence of earlier treatment elsewhere, or absence of overlapping or repeated treatment episodes.

### Methods S2 Order-based endpoints

For each patient and each drug, all qualifying captured orders were treated as a single drug-specific order series. The record span was the last qualifying order date minus the first qualifying order date. A series with one dated order had a span of zero days. No final dosing interval or grace period was added, and the measure did not use a clinical discontinuation event or a time-to-event censoring rule. Interruptions and gaps between recorded orders remained within the span. The measure may combine more than one treatment episode and is not time on treatment, progression-free survival or a validated surrogate for either.

The main descriptive contrast was the difference in median second-drug record span between SG→T-DXd and T-DXd→SG. The second drug differs between these groups, and cohort eligibility requires eventual orders for both drugs. Consequently, this contrast combines drug identity, sequence position, patient selection and unmeasured tumor and treatment characteristics but it is not an estimate of the effect of choosing a treatment sequence.

Secondary measures were the first-drug record span, the sum of the two drug spans and the first-to-last span of the later systemic-therapy block, the covered pathway span from the first ADC order to the last recorded systemic-therapy order, any further systemic-therapy order, the number of distinct later systemic agents, and the post-second-series observation span. Summed and covered pathway spans retain gaps and do not measure continuous exposure. Their interpretation also depends on whether overlapping or repeated episodes occurred.

Further systemic therapy required an order for an agent other than T-DXd or SG after the last order in the second-drug series. Continuing endocrine or HER2-directed agents could satisfy this definition. It does not establish a new treatment line or distinguish death from subsequent care not captured in the database. Prior, intervening and subsequent systemic agents were identified using the executed name-token list and the complete original list must accompany the final reproducibility materials rather than being reconstructed from drug counts.

The inter-drug interval was the first order date of the second drug minus the last order date of the first. The post-second-series observation span was the latest dated record of any type minus the last order date of the second drug. This describes available record coverage, not validated clinical follow-up or vital status. Overall survival was not analyzed because mortality ascertainment, treatment-initiation dates, and follow-up were not established as validated inputs to this analysis. If these inputs are validated, survival from second-ADC initiation could describe prognosis among patients reaching that treatment but it would not estimate the effect of choosing an ADC sequence at the first treatment. Outcomes beginning at the first ADC remain conditioned on a later order for the second drug and exclude patients who never had that order, including those with prolonged first-drug benefit, death or loss of capture.

### Methods S3 Patient characteristics and variables not used

Characteristics were age, recorded sex or gender, race, ethnicity, the number of distinct previously ordered systemic agents, selected metastasis diagnosis indicators. Intervening systemic orders were treatment-pathway measures and are reported in Table S3. C78 codes identified selected visceral metastasis records and C79.3 codes identified brain or cerebral meningeal metastasis records on or before the first ADC order. These are limited code-based indicators where absent codes do not establish absence of metastases, and the C78 and C79.3 families do not encompass every visceral or CNS metastatic site.

Sex, race and ethnicity were obtained from the most recent available demographic record by record date and mapped to PCORnet Common Data Model version 6.0 categories. Self-report status was unknown and the source lacked a validated demographic update timestamp. The sex field may represent gender. Other race could include unknown values. Categories were combined as needed for small-cell suppression, and the ethnicity category remaining after 'not Hispanic' included both Hispanic and other records and it must not be described as exclusively Hispanic.

HER2 status, hormone receptor status, performance status, disease burden, regimen-level line of therapy, treatment response and reasons for stopping were not available as validated analytic variables. Although clinical notes were included in the source, discontinuation reasons were not validated for this analysis. Occurrence of toxicity and attribution of treatment cessation are distinct. Future abstraction should link a documented treatment decision to a specific ADC episode and distinguish progression, toxicity, other reasons, and uncertain or undocumented reasons, with clinician adjudication. Note-derived clinical labels were not used as exposures, outcomes, subgroups or covariates. The manuscript makes no claim that agreement between AI runs validates such labels. No adjusted comparison of sequence groups is retained in the reported analysis. The timing and derivation of age should be documented from the executed source query before submission.

### Methods S4 Order-interval plausibility assessment

The supplied analysis applied an operational eligibility rule requiring at least three orders for a drug or any procedure record from 7 days before the first order through 7 days after the last order. This rule identifies order series with additional recorded activity but does not confirm infusion, administration or completion of treatment. An unrelated procedure or repeated orders for a single intended dose can satisfy the rule. Because SG is ordinarily ordered twice within a 21-day cycle and T-DXd once, a fixed order-count rule has different temporal implications for the drugs.

Among series satisfying that rule and containing at least two orders, the mean interval was calculated as the first-to-last span divided by (number of orders − 1). The cohort summary is the median and IQR of those per-series means, not the median of all individual adjacent intervals. Orders on the same day were not systematically deduplicated. Repeated or rescheduled orders can therefore shorten this statistic without indicating more frequent administration.

The interval summaries were assessed for broad consistency with the drugs' dosing schedules. Agreement at the aggregate level cannot validate individual order identity, administration or endpoint accuracy. Diagnosis-code and supportive-care indicators do not establish adverse-event incidence, severity or attribution. The source comments reported that harmonized laboratory values were inaccessible through the permitted interface. No validated laboratory extract was supplied for this revision. No laboratory-based severity grades are reported. Quantitative safety comparisons and unverified laboratory-monitoring counts are not retained in the revised results.

### Methods S5 Contextual comparison with published evidence

Published sequencing studies provide clinical context in Figure 1 and Table S5. The comparison includes full reports, congress abstracts, and a separately labeled published meta-analysis. Table S5 records populations, endpoints, subgroup denominators, medians, and source qualifications. We did not perform a new systematic review or pool estimates. Cohorts may overlap, and sample sizes must not be summed across studies. ADC-Low subgroup estimates are matched by hormone receptor status and sequence; first-ADC PFI and second-ADC PFS remain distinct endpoints. AURORA medians are not connected because their denominators are not demonstrated to match.

Progression-free survival, real-world progression-free survival, and time to treatment failure differ from first-to-last order spans. No cross-study test, calibration, equivalence assessment, or validation of order spans was performed. Published findings cannot validate the source cohort's medication identity, dates, or clinical endpoints. No effect of AI assistance on extraction accuracy or efficiency was measured.

### Methods S6 Statistical analysis and analysis amendments

Record spans and other continuous measures were summarized using medians and IQRs, with means also reported for counts of distinct later agents. The supplied comparisons used two-sided Mann-Whitney U tests with a tie-corrected normal approximation. This test concerns distributions rather than only their medians. Differences in medians were expressed as SG→T-DXd minus T-DXd→SG, with percentile bootstrap 95% confidence intervals from 5,000 resamples and reported seed 7. The original Python version and random-number generator require confirmation from the executed analysis environment. The editorial review did not independently recompute patient-level bootstrap intervals.

Proportions were summarized with Wilson confidence intervals and between-group differences with Newcombe confidence intervals. Further systemic therapy was compared using a pooled two-proportion z test. Holm adjustment was applied across the six secondary comparisons: first-drug span, summed span, covered pathway span, further systemic therapy, number of later agents and post-second-series observation span. The inter-drug interval was descriptive. No absence-of-difference test was interpreted as equivalence, and the analyses do not support a preferred clinical sequence.

The current record-span analysis amends an earlier approach that applied an order-count filter, added 21 days to the last order and used time-to-event methods. That approach was retired because the filter depended on drug schedule and the event/censoring specification was insufficiently documented. Era-stratified analyses were not retained because dates were shifted. The revised analyses are exploratory; the current endpoint should not be described as prospectively prespecified without a dated amendment establishing that history. Appendix S1 records these amendments without reintroducing withdrawn numerical estimates.

Counts from 1 through 10 and complementary cells are withheld according to the source release restrictions. All reported observations are aggregate. The analyses cannot be independently reproduced from medians, IQRs and group sizes alone; source-environment code, the executed codelists, environment details and permitted output records are required for that purpose.

### Methods S7 Human-Guided Orchestration and Agentic Research Workflow

Muse Spark 1.3 (Meta Superintelligence Labs) operated in a Symbiotic Agent Teaming (SAT) environment, utilizing tools for code execution in the licensed analysis environment; retrieval from PubMed, PubMed Central, Crossref, OpenAlex, and ClinicalTrials.gov; and file editing. The agent wrote and executed cohort, endpoint, and statistical code; performed data-quality checks; retrieved and compared published evidence; generated figures; and drafted manuscript text and comment responses.

Investigators provided Human-Guided Orchestration (HGO) by defining the clinical question and directing the analysis, while acting as an Expert-in-the-Loop (EITL) to review code and evidence, and to decide on changes prompted by coauthor comments and advisory agent critiques. Table 3 in the main manuscript maps these reported stages, architectural responsibilities, and outputs.

End-to-end refers to question formulation through scientific reporting. Workflow outputs and scientific revisions are described as a case study, without an independent comparator or a prespecified AI-performance endpoint. The original duration analysis was amended, and note-derived labels were excluded from analytic variables following EITL review. These were documented scientific decisions rather than measures of agent accuracy. No new agent architecture, measured efficiency gain, autonomous clinical discovery, or independent reproduction is claimed. Execution records support traceability and review and do not resolve limitations in source-data completeness, clinical measurement, or study design. Exact orchestration and execution details should be reported from the original records.

Subsequent editorial review included a multi-dimensional critique, selected source checks, arithmetic verification from reported counts, figure generation, and document production, without access to clinical records or execution of the original cohort queries. The accompanying aggregate audit checks only disclosed counts, Wilson and Newcombe intervals, and the pooled two-proportion test. It cannot verify individual dates, medians, IQRs, bootstrap CIs, Mann–Whitney tests, or the original Holm calculations.

**Table S1** Cohort ascertainment

| Stage | Reported count | Interpretation |
|---|---|---|
| Database release | 4,950,000 | Patients in the June 4, 2026 release |
| C50-family diagnosis code | 156,051 | Any breast cancer code; not an ADC-eligible denominator |
| Reported analytic cohort | 106 | C50-family diagnosis, orders assigned to both drugs, and a resolvable first-order sequence after date checks |
| T-DXd ordered first | 73 | First observed T-DXd order predates first SG order |
| SG ordered first | 33 | First observed SG order predates first T-DXd order |

Intermediate exclusion cells are suppressed. Author verification of drug names, brand-name capture, and shifted-date exclusions required no change to these cohort counts. The release contains patients with unknown stage, receptor status, treatment eligibility, and observation history.

**Table S2** Operational definitions used for the reported aggregates

| Variable | Executed rule and interpretation |
|---|---|
| Breast cancer | Diagnosis code beginning C50; ICD-10-CM diagnosis records; Any date |
| T-DXd order | Name contains "trastuzumab" and "deruxtecan"; Source medication name, case-insensitive; all order statuses; Order start date, or order record date when missing |
| SG order | Name contains "sacituzumab" or "govitecan"; Source medication name, case-insensitive; all order statuses; Order start date, or order record date when missing |
| Treatment order | Earlier first qualifying order date; ties excluded; Derived; Within patient |
| Selected C78 codes | Diagnosis code beginning C78; ICD-10-CM; On or before the first ADC order |
| Selected C79.3 codes | Diagnosis code beginning C79.3; ICD-10-CM; On or before the first ADC order |
| Prior and intervening systemic agents | Original executed name-token list, not supplied; Source medication name; Before the first ADC order; between the drugs |
| Further systemic therapy | Original executed name-token list, including trastuzumab emtansine and datopotamab deruxtecan; Source medication name; After the last order of the second drug |
| Order-density criterion | At least 3 orders, or a procedure record of any type; Medication orders; procedure records; Procedure from 7 days before the first order through 7 days after the last |
| Last recorded activity | Latest dated record of any type; All record types; Within patient |
| Sex, race, ethnicity | Most recent demographic record, by record date; PCORnet Common Data Model version 6.0 categories in the analysis data; Not applicable |

These are the executed rules. Drug-name verification found no incorrect drug assignment or brand-name omissions affecting the cohort; sacituzumab tirumotecan was absent, and no clinical-trial participant data were included in the primary analysis. No shifted-date exclusions affected the cohort. The full nonstudy-agent codelist remains an author completion item. Planned orders and untied placeholder dates remain measurement limitations; name verification does not establish administration.

**Table S3** Full reported record span and other systemic therapy comparisons

| Measure | T-DXd→SG (n = 73) | SG→T-DXd (n = 33) | Difference (95% CI) | P value | Holm P value |
|---|---|---|---|---|---|
| Second-drug record span, days, median (IQR) | 63 (29–116) | 49 (21–126) | −14 (−41 to 21) | 0.42 | — |
| First-drug record span, days, median (IQR) | 141 (71–334) | 189 (115–419) | 48 (−33 to 200) | 0.09 | 0.54 |
| Interval between the drugs, days, median (IQR) | 64 (30–161) | 40 (14–126) | — | — | — |
| Other systemic therapy between the drugs, No. (%) | 32 (44) | Suppressed | — | — | — |
| Summed record span, days, median (IQR) | 287 (168–510) | 393 (196–590) | 106 (−67 to 315) | 0.17 | 0.83 |
| Covered pathway span, days, median (IQR) | 426 (258–714) | 514 (316–756) | 88 (−133 to 278) | 0.35 | >0.99 |
| Further systemic therapy, No. (%) | 47 (64) | 17 (52) | −13 (−32 to 7) | 0.21 | 0.84 |
| Distinct later agents, mean (median) | 1.45 (1) | 1.24 (1) | — | 0.41 | >0.99 |
| Recorded observation after second drug, days, median | 154 | 184 | — | 0.62 | >0.99 |

Differences are SG→T-DXd minus T-DXd→SG. Median differences are in days; the proportion difference is in percentage points. P values are retained from the supplied analysis, with 0.089 displayed as 0.09. Reported Holm-adjusted values are retained; unrounded source P values were unavailable for independent verification. The main second-drug comparison is not part of the six-comparison secondary family. Intervening systemic therapy is descriptive, was not tested, and is not part of the six-comparison secondary family. Dashes denote not estimated or not applicable. Observation after the second drug has no IQR in the supplied aggregates. Summed spans can include gaps and overlaps and are not an upper bound on true treatment exposure.

**Table S4** Order interval summary

| Measure | T-DXd | SG |
|---|---|---|
| Per-series mean interval, days, median (IQR) | 21.7 (11.0–28.2) | 12.0 (7.0–17.0) |

The operational density criterion and formula are described in Methods S4. A record series is not an administration-confirmed course. No eligible-series count, excluded complement, toxicity percentage, or laboratory-monitoring count is disclosed. Labels describe every-21-day T-DXd dosing and SG dosing on days 1 and 8 of a 21-day cycle.[17,18]

**Table S5** Published sequencing studies and source qualifications

| Study / reference | Population and sample size | Endpoint | First / second medians, months | Qualification |
|---|---|---|---|---|
| ADC-Low[7] | HER2-low MBC; HR+ T-DXd→SG n=56; HR− SG→T-DXd n=100 | PFI1; PFS2 | HR+: 2.7 / 2.3; HR−: 4.9 / 3.2 | Matched receptor subgroups. Overall-sequence PFS2 was 2.2 (T-DXd→SG, n=64) and 3.1 (SG→T-DXd, n=115); those estimates are not paired with subgroup PFI1. Patients alive and progression-free at cutoff were excluded. |
| ADC-Low-Europe[20] | HER2-low MBC; N=331 overall; exact sequence-subgroup n not reported | PFI1; PFS2 | HR+ T-DXd→SG: 6.3 / 2.4; HR− SG→T-DXd: 5.4 / 2.7 | Congress abstract. Receptor status closely tracks sequence; PFI1 and PFS2 differ. Possible overlap with ADC-Low. |
| City of Hope[21] | HER2-low MBC; T-DXd→SG n=70; SG→T-DXd n=30 | PFS | T-DXd→SG: 6.8 / 2.5; SG→T-DXd: 5.6 / 2.3 | Congress abstract; single center. Full endpoint ascertainment details are not provided in the abstract. |
| Portuguese national study[22] | HER2-low advanced breast cancer; n=64; 14 centers | PFS | Both sequences combined: 8.9 / 2.7 | Congress abstract. Sequence-specific medians not reported. T-DXd first n=34; SG first n=30. |
| Huppert et al[6] | HER2-low MBC; HR+ n=56; HR− SG→T-DXd n=25 | TTF | HR+ T-DXd→SG (n=32): 5.3 / 2.1; HR+ SG→T-DXd (n=24): 6.3 / 3.6; HR− SG→T-DXd (n=25): 7.5 / 2.8 | Five-center retrospective cohort. TTF includes discontinuation for any reason. HR− T-DXd→SG (n=3) medians were undetermined. |
| Flatiron: after T-DXd[9] | HR+/HER2− MBC; SG subgroup n not reported in retrieved abstract; N=793 overall | rwPFS | SG after T-DXd: second 2.6; first not reported | Next-line treatment after T-DXd, not a two-sequence comparison. Overall N includes other regimens and subtypes. |
| Flatiron: T-DXd[8] | TNBC with prior SG, n=58; N=1,490 overall | rwPFS | T-DXd after prior SG: second 3.4; first not reported | Prior SG at any time, not necessarily the immediately preceding line. No-prior-SG comparator n=61 had rwPFS 5.7 months; it is not an ADC1 estimate. |

**Table S5 (continued)**

| Study / reference | Population and sample size | Endpoint | First / second medians, months | Qualification |
|---|---|---|---|---|
| AURORA US Network[23] | Mixed MBC subtypes/ADCs; n=72 with ≥1 ADC; n=22 with ≥2 ADCs | Treatment duration | First: 141 days (4.6 months); second: 81 days (2.7 months) | Congress abstract. First-ADC median denominator unstated; includes T-DM1 and other ADCs. Estimates are shown unconnected, not as paired durations in the n=22 subgroup. |
| Meta-analysis[24] | Seven of 17 studies in PFS2 synthesis; T-DXd→SG n=228; SG→T-DXd n=246 | Pooled PFS2 | T-DXd→SG: second 2.8; SG→T-DXd: second 3.3 | Congress abstract; reconstructed curves; $I^2$=70.9%. May overlap individual cohorts. First medians not reported. Internally inconsistent CI for 2.8-month median is omitted. |
| Mai et al 2025[10] | 85 patients with metastatic breast cancer; retrospective | rwPFS | Second-ADC rwPFS was shorter than first-ADC rwPFS in 75.2% of patients. | Not plotted: order-specific medians omitted because the original evidence review identified inconsistent reporting. |
| This study | C50-coded breast cancer; T-DXd→SG n=73; SG→T-DXd n=33 | Medication-order span | T-DXd→SG: 4.6 / 2.1; SG→T-DXd: 6.2 / 1.6 | First-to-last order interval; omits final cycle. Stage and receptor status not validated. Not PFS, TTF, or verified administration duration. |

Selected evidence provides clinical context; this is not a new systematic review or meta-analysis. Only the explicitly labeled published meta-analysis row is pooled. The sources may overlap. Medians do not have comparable clinical meaning across endpoints. MBC, metastatic breast cancer; HR, hormone receptor; HER2, human epidermal growth factor receptor 2; PFI, progression-free interval; PFS, progression-free survival; rwPFS, real-world PFS; TTF, time to treatment failure; TNBC, triple-negative breast cancer; NR, not reported. Reference numbers correspond to the main manuscript.

**Table S6** Data quality findings and unresolved limits

| Domain | Finding | Interpretation or unresolved evidence |
|---|---|---|
| Drug identity | Drug names verified with no changes | No tirumotecan records or brand-name omissions affecting the cohort; no clinical-trial participant data in the primary analysis |
| Date conformance | Valid-calendar check excluded no patients | Valid dates can still be placeholders; cross-table shift consistency is unconfirmed |
| Temporal boundaries | Shifted-date exclusions checked | Author verification found none affecting the analytic cohort; results unchanged |
| Order uniqueness | No systematic deduplication | Short T-DXd intervals may reflect repeated or revised orders |
| Observation | Latest recorded activity available | Not validated follow-up or a common clinical censoring boundary |
| Clinical variables | Stage and biomarkers not validated | No subtype-specific or adjusted sequence-effect claim |
| Clinical validation | No physician reference standard established | Plausibility assessments do not yield positive predictive value |
| Analysis provenance | Only documents supplied for this revision | Original code, unrounded statistics, codelists and run manifest needed |

**Table S7** STROBE and RECORD reporting map

| Reporting item | Location and reporting status |
|---|---|
| Title and design | Abstract and Methods, Design and data source (main pp. 2–3). Retrospective descriptive EHR study; analyses are unadjusted. The human-guided agentic workflow is reported as a worked demonstration. |
| Setting and dates | Methods, Design and data source (main pp. 3–4); Methods S1 (supplement pp. 1–2). Database release and overall coverage are described. The study-specific date range remains bracketed; outside-system capture descriptions require reconciliation. |
| Eligibility and selection | Methods, Cohort and medication orders (main p. 4); Methods S1 and Tables S1–S2 (supplement pp. 1–2, 7–8). C50-family diagnosis, orders for both drugs, and resolvable sequence define eligibility; small intermediate exclusion counts are suppressed. |
| Exposure algorithms and codelists | Methods, Cohort and medication orders (main p. 4); Methods S1–S2 and Table S2 (supplement pp. 1–2, 8). Study-drug matching, order-status retention, date fallback, and sequence rules are reported. The complete executed codelist for other systemic agents is not supplied. |
| Validation of algorithms | Methods, Characteristics and data quality (main p. 5); Methods S1, S4 and Table S6 (supplement pp. 1–4, 13). Drug names and exclusion impact were verified. Administration and clinical endpoints were not validated against an independent clinical reference standard. |
| Variables and outcomes | Methods, Endpoints and interpretation and Characteristics and data quality (main pp. 4–5); Methods S2–S4 and Tables S2–S4 (supplement pp. 2–4, 8–10). Order spans, later therapy, characteristics, and interval measures are defined; spans are not validated clinical outcomes. |
| Bias and confounding | Methods and Discussion (main pp. 4–5, 8–9); Methods S2–S3 (supplement pp. 2–3). Selection on orders for both ADCs, incomplete capture, unvalidated clinical covariates, and non-equivalence of sequence groups are acknowledged. Comparisons are unadjusted and do not identify a preferred sequence. |
| Study size | Methods, Statistical analysis and Results, Selected cohort (main pp. 5–6); Table S1 (supplement p. 7). The available release yielded 106 eligible patients, with sequence groups of 73 and 33. No prospective sample-size calculation is asserted. |
| Missingness and linkage | Methods, Design and data source (main pp. 3–4); Methods S1, S3 and Table S6 (supplement pp. 1–3, 13). Database-level claims linkage is described. Cohort-specific linkage completeness, variable-level missingness, and cross-table date-shift consistency are not established. |

**Table S7 (continued)**

| Reporting item | Location and reporting status |
|---|---|
| Statistical methods | Methods, Statistical analysis (main p. 5); Methods S6 and Table S3 (supplement pp. 4–5, 9). Mann–Whitney tests, bootstrap median-difference intervals, Wilson and Newcombe intervals, a pooled proportion test, and Holm adjustment for six secondary comparisons are specified. |
| Participant characteristics | Results, Selected cohort and Table 1 (main pp. 6, 13); Methods S3 (supplement p. 3). Recorded demographics, prior-agent counts, and selected diagnosis indicators are reported with suppression. Age timing is unresolved; receptor status and treatment line are not validated. |
| Outcome estimates | Results, Record spans and later orders; Table 2 and Figures 1–2 (main pp. 6–7, 14, 16–17); Tables S3–S5 (supplement pp. 9–12). Estimates, uncertainty, and contextual evidence are reported. Intervening systemic therapy is reported descriptively in Table S3. |
| Other analyses and changes | Methods, Human-Guided Orchestration and Agentic Research Workflow (main pp. 5–6); Methods S4, S6–S7 and Appendix S1 (supplement pp. 3–6). Order-interval assessment and amendments are described; filtered duration, era, and weighted analyses were withdrawn. |
| Limitations and generalizability | Discussion (main pp. 8–9); Methods S1–S5 and Table S6 (supplement pp. 1–4, 13). Selection, measurement, clinical validation, care capture, sample size, and disclosure limits are described. Clinical sequence effects and comparative AI performance are not established. |
| Funding and conflicts | Funding and support (main p. 9). No specific grant funding is reported; Meta provided Muse Spark compute. Author competing-interest declarations and the funder's role beyond compute provision are not reported in the current manuscript. |
| Protocol data and code access | Data and code availability (main p. 9); Methods S6–S7 and Appendices S1–S3 (supplement pp. 4–6). Licensed-data restrictions, amendments, and limited aggregate-audit materials are described. No public repository DOI is claimed. |
| Ethics | Methods, Ethics and reporting (main p. 6). Bluebonnet is the author-confirmed IRB of record; the main-text IRB name needs alignment. Licensed deidentified records and privacy suppression are described. Bluebonnet Ethical Review served as the institutional review board of record. Its determination was that this activity does not meet the regulatory definition of human subjects research under the U.S. Department of Health and Human Services regulations at 45 CFR 46.102. |

Page numbers refer to the current manuscript and supplement PDFs. This map identifies where reporting items are addressed and where information remains incomplete. It does not establish full compliance with the STROBE and RECORD checklists.
